# Early diagnosis of oral cancer and lesions in Fanconi anemia patients: a prospective and longitudinal study using saliva and plasma

**DOI:** 10.1101/2023.01.03.22284070

**Authors:** Ricardo Errazquin, Estela Carrasco, Sonia Del Marro, Anna Suñol, Jorge Peral, Jessica Ortiz, Juan Carlos Rubio, Carmen Segrelles, Marta Dueñas, Alicia Garrido-Aranda, Martina Alvarez, Cristina Belendez, Judith Balmaña, Ramon Garcia-Escudero

## Abstract

Fanconi anemia (FA) patients display an exacerbated risk of oral squamous cell carcinoma (OSCC) and precursor lesions at young ages, mainly at the oral cavity. As patients have defects in DNA repair mechanisms, standard-of-care treatments to OSCC such as radiotherapy and chemotherapy give rise to severe toxicities. New methods for early diagnosis are urgently necessary to allow treatments in early disease stages and achieve better clinical outcomes. We have conducted a prospective, longitudinal study whereby liquid biopsies from sixteen lesion/tumor-free patients were analyzed for the presence of mutations in cancer genes. DNA from saliva and plasma were sequentially collected and deep-sequenced, and the clinical evolution followed during a median time of around 2 years. In 9/16 FA patients we detected mutations in cancer genes (mainly *TP53*) with molecular allele frequencies (MAF) down to 0.07 %. Importantly, all patients having mutations and clinical follow-up data after mutation detection (n=6) developed oral precursor lesions or OSCC. Lead-time between mutation detection and tumor diagnosis ranged from 23 to 630 days. Strikingly, FA patients without mutations display significantly lower risk of developing precursor lesions or OSCC. Therefore, our diagnostic approach could help to stratify FA patients into risk groups, which would allow closer surveillance for OSCC or precursor lesions.

## INTRODUCTION

Head and neck squamous cell carcinoma (HNSCC) arises in the mucosal linings of the upper aerodigestive tract, which includes the oral cavity, larynx and pharynx [1]. Carcinomas in the oral cavity can evolve from visible oral potentially malignant lesions (OPMLs) such as leukoplakias and erytroplakias (white or red patches, respectively) [2]. The clinical outcome of OPMLs is variable, with many lesions that regress. Risk assessment methods are based on histopathology analysis from tissue biopsies, but they can fail as some low-risk lesions could become malignant [3]. What is needed is the development of more accurate prediction methods, possibly including genetic analysis such as loss of heterozygosity (LOH) [4]. Furthermore, lesions can be treated with surgery [5], but even when margins are negative, recurrence rate is high, which demonstrates that active treatment does not always prevent OSCC [6]. These recurrent tumors might arise from mutant precancerous fields that display normal histology [7]. Occasionally, patients display large or multifocal lesions that hamper surgical removal. Therefore, close surveillance of OPMLs might be a better option until new active therapies are developed [8]. On the other hand, most patients present with tumors *de novo*, without previous clinical symptoms. These tumors originate from invisible premalignant cells, possibly within precancerous fields [7]. Currently, there are no tools to detect such cells before carcinoma development, thus hampering an appropriate clinical management of OSCC patients.

Risk factors for OSCC and OPMLs include alcohol and tobacco consumption, as well as genetic predisposition. Fanconi anemia (FA) is a genetic syndrome with a propensity to congenital malformations, bone marrow failure (BMF), and cancer [9, 10]. Most FA children develop BMF that is treated with hematopoietic stem cell transplantation (HSCT), which significantly improves life expectancy. Current advances in a gene therapy clinical trial might provide an alternative to HSCT in the near future to FA children potentially developing BMF [11]. Sadly, HNSCC propensity is the most important health challenge nowadays in FA patients, with incidences 500 times higher than the general population [12]. OSCC in FA is associated with a high mortality rate. Median age at diagnosis is around 25-31 years old, half the age of standard patients. Pathogenic mutations have been reported in 23 different genes that give rise to Fanconi or Fanconi-like clinical phenotype [9, 10]. A hallmark feature of FA is the inability to repair DNA interstrand cross-links (ICLs). FA proteins repair ICLs in a common cellular pathway known as the FA pathway or FA/BRCA pathway. HNSCC in FA is difficult to treat, as patients cannot tolerate current therapies, which include ionizing radiation and platinum-based chemotherapies [13]. In addition, most tumors are detected at advanced stages. Altogether, these events have made cancer the principal cause of early mortality in adults with FA. Therefore, the discovery of non-genotoxic and efficient treatment opportunities for FA patients is of utmost importance

FA individuals develop OPMLs very frequently, mainly leukoplakias, at much higher rates than the general population: 9-12% versus 1-2%, respectively [14-16]. Furthermore, average age at presentation is much younger: 15-16.5 years old in FA patients versus 45-54 years old in non-FA patients. Oral lesions in FA are clinically challenging as they frequently involve large areas and appear multifocally. Patients display consecutive lesions, which recur frequently and can progress to malignant carcinomas. Importantly, FA individuals with leukoplakias are at higher risk of OSCC. Close surveillance is important and should begin at pediatric ages, especially after HSCT. Therefore, there is an urgent clinical need for new technologies that enable accurate detection of premalignant cells. Due to close and repeated examination, a noninvasive technique would be desirable, as tissue biopsies are a burden for the patient.

Liquid biopsies (LBs) constitute a non-invasive source of patient biological material that can help to diagnose pathologies such as cancer. LBs can be collected easily and repeatedly during pathologies that require close surveillance. Methods to detect cancer biomarkers in LBs, such as tumor DNA, have substantially improved over the last decade. Thus, some technologies based on next generation sequencing (NGS) can detect tumor DNA molecules with limit of detection (LOD) of <0.1%. This high sensitivity has been particularly useful when analyzing circulating tumor DNA (ctDNA) in plasma. Detection of ctDNA provides real-time knowledge of cancer, starting with screening before it is clinically apparent [17] as well as during all stages of cancer treatment [18]. Importantly, LBs have already entered clinical practice, guiding treatment in a subset of lung tumors. Besides plasma, another LB with potential utility for cancer research is saliva. In HNSCC patients, the existence of tumor cells and tumor-derived DNA (tDNA) in saliva has been reported [19]. As a tumor grows, it releases tDNA into saliva so that mutations found in tDNA are equal to those of the primary tumor [20, 21]. Therefore, tDNA could be used as a surrogate for a tissue biopsy. Indeed, Wang et al reported that saliva and plasma analysis allowed detection of somatic mutations and human papillomavirus (HPV) in 96% of HNSCC patients (100% in oral cavity) [21], showing that the evaluation of plasma can complement that of saliva. To our knowledge, the utility of tumor DNA detection in saliva and plasma for early diagnosis of OSCC in high-risk patients has not been tested so far.

Here, we present a proof-of-principle study whereby NGS-based, sensitive detection of cancer DNA mutations in saliva and plasma from FA patients could help to detect OPMLs and/or OSCC before clinical diagnosis. LBs from patients were repeatedly collected in a prospective and longitudinal clinical study of around 2 years. Results demonstrated that our biomarker approach could help to identify FA patients at very high risk of OPMLs and OSCC.

## MATERIALS AND METHODS

### Study design and patient cohort

This is a multi-centre non-interventional prospective longitudinal cohort clinical study. Fanconi anemia patients with no previous confirmed evidence of the development of solid or blood malignancies were considered eligible. Patients with myelodysplastic syndrome were excluded, as well as those with major health issues. Sixteen Spanish patients were recruited at the Hospital Universitario Gregorio Marañon (Madrid, Spain) and the Hospital Universitario Vall d’
sHebron (Barcelona, Spain) between November 2018 and December 2021. The primary objective was the detection of mutations in cancer genes in DNA from liquid biopsy samples prior to clinical diagnosis of lesions or carcinomas in the head and neck. We included HSCT patients older than 14 years old (n=11), and non-HSCT patients older than 25 years old (n=5). Clinical follow-up at the Oral Dentistry Department has been conducted as part of the clinical surveillance. All patients gave their consent by signing a form appropriate to the study, approved by the Ethics Committee for Clinical Research of the hospitals involved. Saliva and whole blood were collected from each patient at least once during the duration of the project, and yearly whenever possible. Sample/patient IDs are not known to anyone outside the research group.

### Saliva and plasma collection and DNA purification

Saliva samples were collected as oral rinses. Patients were asked to swish 10 ml of 0.9% sodium chloride in their mouths for 10 to 15 s before spitting into the collection tube. Venous blood (4-10 ml) was collected by standard phlebotomy techniques in EDTA tubes and maintained at 4°C until further processing. Plasma was obtained from blood within 4 hours of phlebotomy upon two sequential centrifugations: first at 1200 x g for 7 min at 4ºC; and second, 14000 x g for 10 min at 4ºC. In some instances, the buffy coat layer was recovered during blood processing, lysed using BD FACS Lysing Solution (Thermo Fisher Scientific, Waltham, MA, USA), centrifuged at 3000 rpm for 10 minutes, and stored at -80°C. Saliva genomic DNA (gDNA) was extracted from 400 μl of oral rinses using MagMAX Saliva gDNA Isolation Kit (Thermo Fisher Scientific, Waltham, MA, USA). Plasma circulating cell-free DNA (cfDNA) was extracted from 4 ml of plasma using MagMAX Cell-Free Total Nucleic Acid Isolation Kit (Thermo Fisher Scientific, Waltham, MA, USA). Leukocyte DNA was extracted using DNeasy Blood & Tissue Kit (Qiagen). DNA was quantified using Qubit Fluorometric Quantification (Thermo Fisher Scientific, Waltham, MA, USA).

### Library preparation and deep-sequencing

Libraries for next generation sequencing (NGS) were constructed from 10-50 ng of DNA using the Oncomine™ Pan-Cancer Cell-Free Assay (OPA) (Thermo Fisher Scientific, Waltham, MA, USA) and quantified using Ion Library TaqMan™ Quantification Kit (Thermo Fisher Scientific, Waltham, MA, USA). Templating and sequencing were performed using the Ion 540™ sequencing chip on the Ion Chef and Ion S5 XL systems (Thermo Fisher Scientific, Waltham, MA, USA), according to the manufacturer’s instructions. Although OPA is designed to detect some gene fusions from RNA, we did not analyze them.

### Sequencing analysis

Sequencing raw data were analyzed on the Torrent Server™, reads aligned to the reference human genome (hg19), and variant analysis performed using the Ion Reporter™ Analysis Server. Quality control was done manually for every sample based on the following filters: number of *mapped reads* per sample > 10,000,000; and *on-target reads* > 90%. For samples with missing data on these filters, we filtered out those with *median molecular coverage* <500. *Median molecular coverage* is the median number of individual interrogated DNA molecules across targets. We always require two independent molecular families to identify a variant. Variant calling was performed with the Ion Reporter Analysis Software v.5.18 using the Oncomine TagSeq Pan-Cancer Liquid Biopsy w.2.5-Single Sample workflow and the predefined filter chains Variant Matrix Summary (5.18) and Oncomine Extended (5.18). Only variants with at least 2 molecular counts and with a molecular allele frequency (MAF) of >0.065% were included in the analysis. MAF is the mutant allele proportion among the total molecular number (consisting of both wild-type and mutated sequences) expressed as a percentage. For copy number variants (CNVs) we included those with a ratio above 1.15 and with a median absolute pairwise difference (MAPD) below 0.4. Using these thresholds, we did not detect CNVs in our LB samples. Known germline variants with allelic frequency above 35% or single nucleotide polymorphisms were not included in the analysis. Leucocyte DNA from some FA patients was similarly sequenced and analyzed.

### Survival Curves

Kaplan-Meier survival curves were obtained with Prism software v.9.0 (Graphpad Software, Inc., www.graphpad.com). Statistical significance of survival between genotypes was calculated with the log-rank test yielding a p-value.

## RESULTS

### Patients’ characteristics

A total of 16 Fanconi anemia patients from two clinical centers in Spain (Hospital Vall d’Hebron in Barcelona and Hospital Gregorio Marañon in Madrid) were included in the study. These patients haven been routinely checked each year at the Departments of Hematology, Maxillofacial, and Hereditary Cancer. At the time of enrollment, patients were lesion/cancer-free at the oral cavity. We included HSCT patients older than 14 years old (n=11), and non-HSCT patients older than 25 years old (n=5). Patient characteristics, clinical follow-up times, lesions and cancer outcomes are detailed in Table 1 and Supplementary Table 1.

**Table 1.** Fanconi anemia patient cohort characteristics

| **Characteristics** | **Fanconi anemia patients (n=16)**<br>**n (%)** |
| --- | --- |
| Age (years) |  |
| Median | 27 |
| Range | 14-46 |
| Sex (n) |  |
| Female | 9 (56) |
| Male | 7 (44) |
| Patients with HSCT | 11 (69) |
| Median years from HSCT to study launch date | 8.4 |
| GVHD | 4 (24) |
| Follow-up time (days) |  |
| Median | 719 |
| Range | 454-1118 |
| Patients with lesions/tumors |  |
| Oral potentially malignant lesion | 6 (35) |
| Oral Squamous Cell Carcinoma | 2 (12) |

Abbreviations: HSCT: hematopoietic stem cell transplantation; GVHD: graft-versus-host-disease.

### Liquid biopsies collected

Saliva and/or plasma were collected at different time points during the longitudinal study (n=50) (Fig. 1). DNA was purified and quantified from 42 saliva and 32 plasma samples with dedicated methods (see M&M). As expected, the amount of DNA was much higher in saliva than plasma (Fig. 2A). Patient genetic material in saliva arose from desquamated keratinocytes during oral rinses, and can potentially contain precancerous or tumor DNA (tDNA). In plasma, purified DNA is cell-free DNA (cfDNA) which normally exists in low quantities in tumor-free patients.

**Figure 1.**
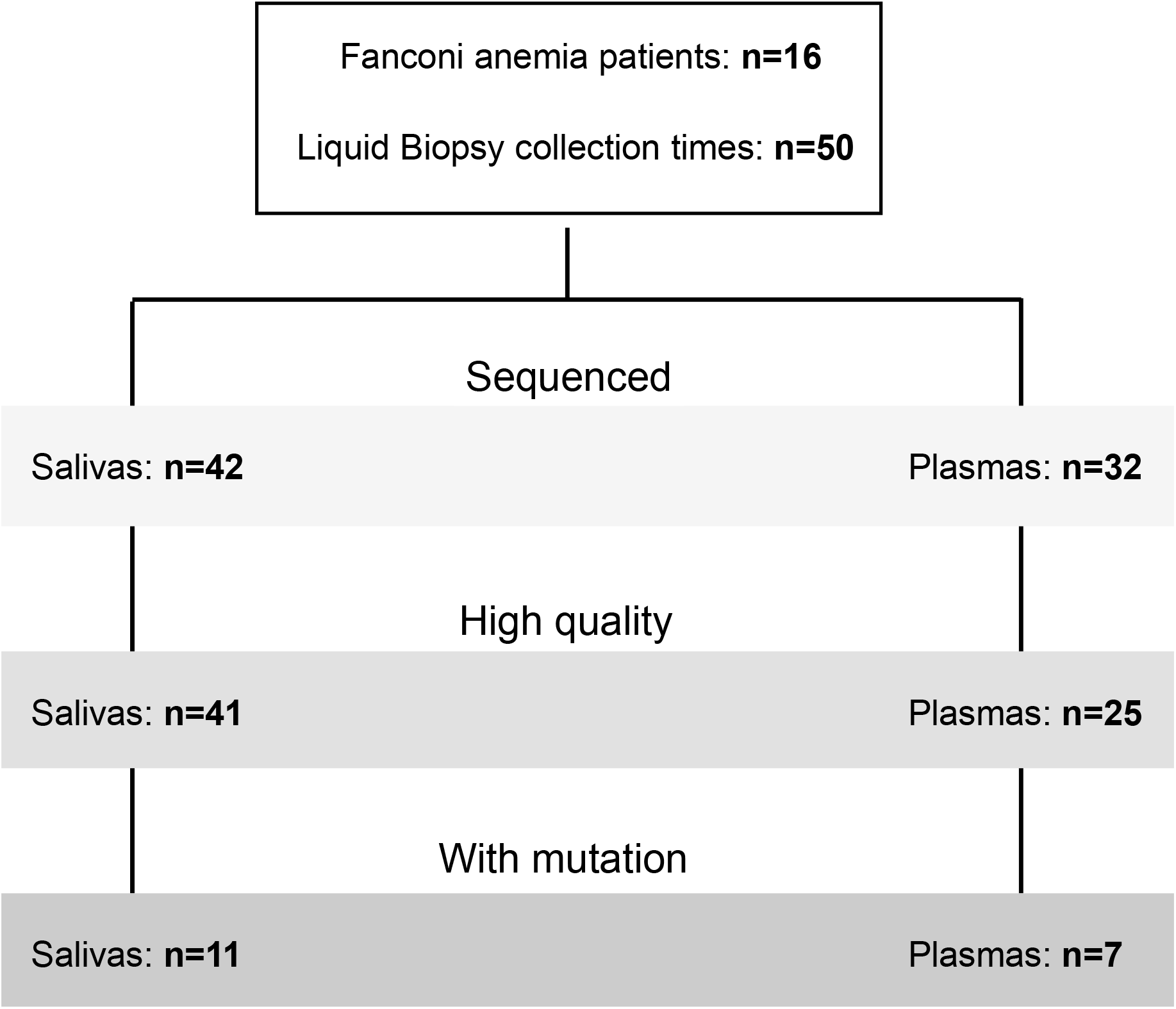
Liquid biopsies collected. A total of 50 collection times of liquid biopsies from 16 FA patients existed during the clinical study. A subset of samples was deep-sequenced with the Oncomine™ Pan-Cancer Cell-Free Assay. Only high-quality sequencing runs were analyzed for the presence of cancer mutations.

**Figure 2.**
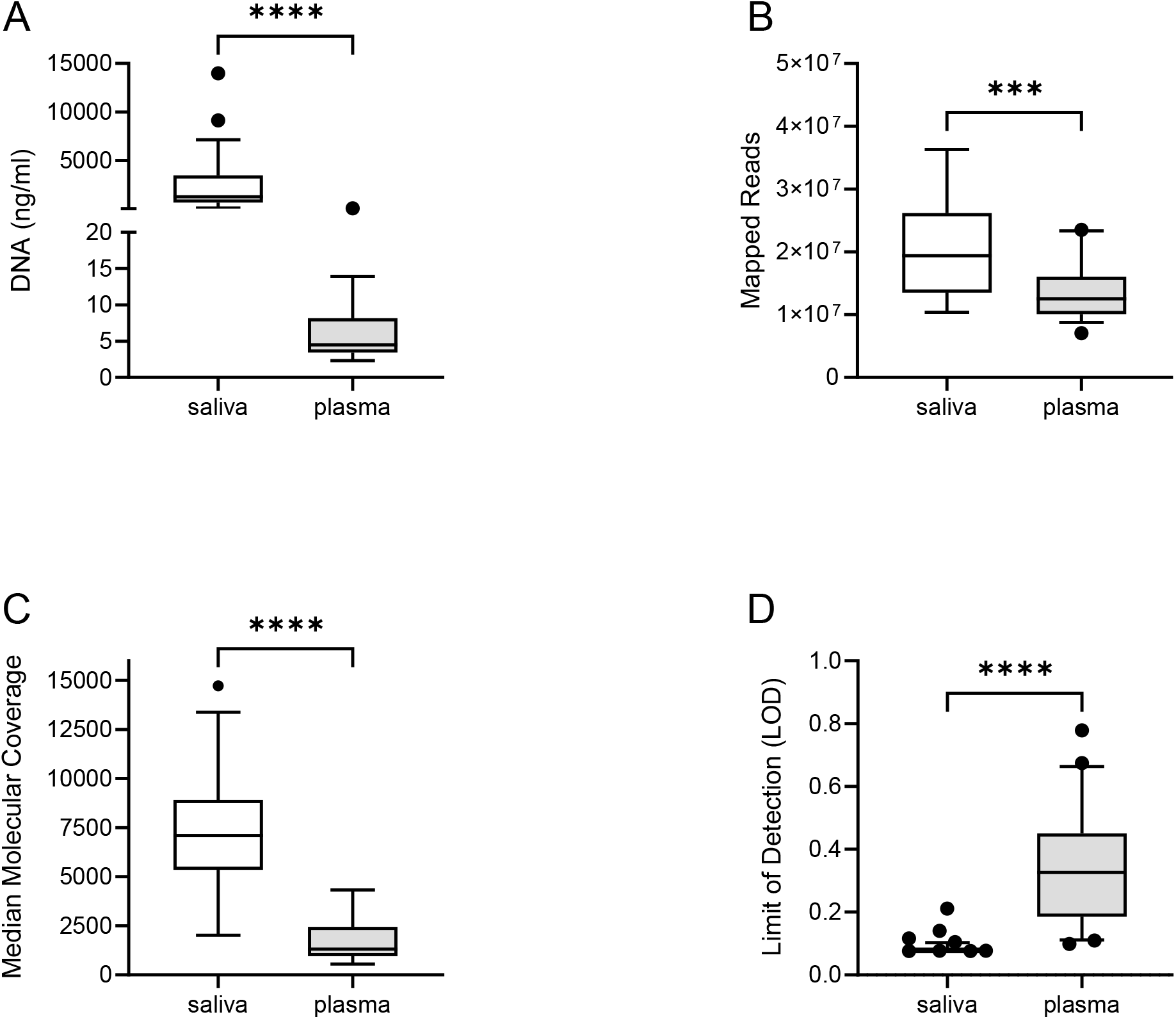
Sequencing performance in saliva and plasma. **A)** The concentration of DNA purified from saliva was significantly higher than that obtained in plasma, as expected. The total number of mapped reads **(B)** and the median molecular coverage **(C)** showed a better sequencing performance of saliva versus plasma. Consequently, the median limit of detection (LOD %) across targets was lower in saliva **(D)**, thus allowing detection of mutations at reduced frequencies in saliva more likely. **: p-value<0.01; ****: p-value<0.0001.

### Sequencing performance in saliva and plasma

DNA from LB was utilized for library preparation and quantification using the Oncomine™ Pan-Cancer Cell-Free Assay (OPA). OPA analysis allows detection of molecular aberrations in 52 genes frequently mutated in solid tumors (Supplementary Table 2). Deep-sequencing was performed using the Ion Chef™ and Ion S5 XL systems (Thermo Fisher Scientific, Waltham, MA, USA). Initial analysis of performance demonstrated that sequencing was optimal for most of the LB samples (Fig. 1), mainly in saliva samples. The number of mapped reads and median molecular coverage (median number of individual interrogated DNA molecules across targets) allowed limit of detection (LOD) values down to 0.07% in saliva and 0.09% in plasma (Fig. 2B-D). LOD values were more variable in plasma samples as the DNA yield obtained was below 20 ng in most cases (Supplementary Table 3).

### Mutations found in liquid biopsies

Although the OPA assay is designed to detect copy-number variants (CNVs) in 12/52 genes (Supplementary Table 2), we did not find CNVs in any LB sample. Importantly, we found non-synonymous small variants or mutations (SNVs, MNVs and INDELs) in 15 samples from 9 patients (Supplementary Table 4). As mutations are found at a low MAF, we could not discard the possibility that they arose from small populations of leucocytes via a selection process such as clonal hematopoiesis (CH). Therefore, we performed the OPA assay using leucocyte DNA obtained from the blood of some of these patients (Supplementary Table 3 and 4). We discarded one sample because of poor sequence quality (FA452) (see M&M). We found a mutation in *IDH2* that was also detected in the saliva of patient FA531. Considering that the MAF observed was below 0.1% in both cases, we suggest that this a mutation associated to CH and not to a lesion/SCC in the oral cavity. Also, we detected a mutation in *PDGFRA* that was not found in the LBs from FA145, so we did not discard any mutation in this patient.

Molecular allele frequencies (MAF %) of mutations observed were similar between saliva and plasma, with minimum values around 0.07% (Fig. 3A). A majority of mutant samples displayed only 1 mutation, but 2 samples had more than 1 mutation (Fig. 3B). Interestingly, most mutations were mapped in the *TP53* gene (Fig. 3C), in line with previous reports showing the high frequency of *TP53* mutations in HNSCC from FA patients [22, 23].

**Figure 3.**
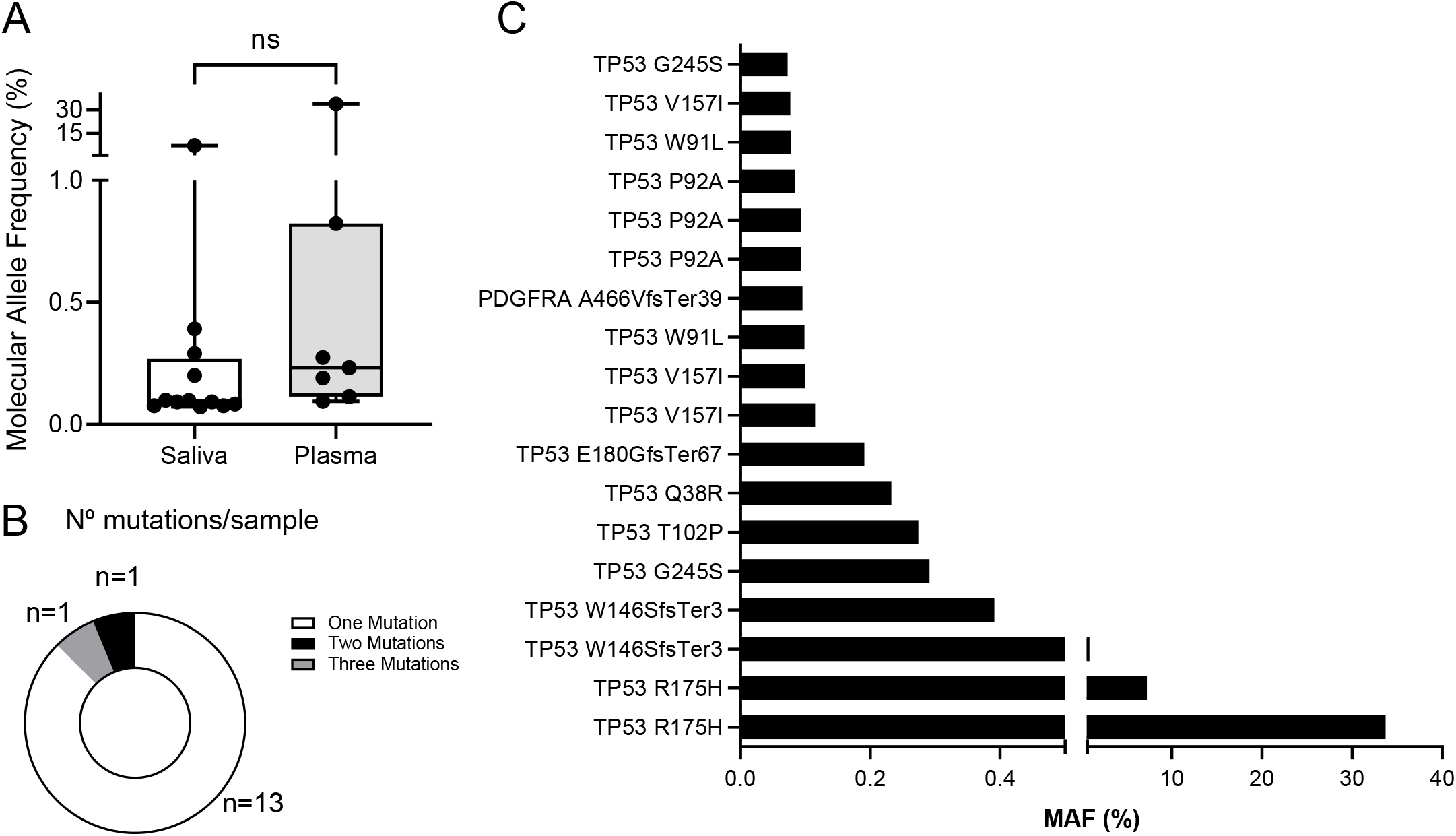
Mutations found in saliva and plasma. **A)** A total number of 18 mutations (SNVs, MNVs and INDELs) were found in 15 samples, with similar molecular allele frequencies (MAF %) between saliva and plasma. **B)** Number of mutations per sample. Samples normally had single mutations (n=13), although multiple concurrent mutations were also found. **C)** Molecular allele frequencies (MAF%) of mutations found. *TP53* was the most frequently mutated gene (95% of samples).

### Association between cancer mutations and clinical outcome

Fourteen FA patients from our cohort had clinical follow-up after collection of at least one LB. The median follow-up time from first LB collection was 719 days. There were 7/14 patients who developed lesions or carcinomas in the oral region, and 7/14 patients who did not (Fig. 4). Most lesions were leukoplakia in the oral cavity (Supplementary Table 5). Two patients had oral squamous cell carcinomas that were subsequently treated with surgery: FA531 and FA145.

**Figure 4.**
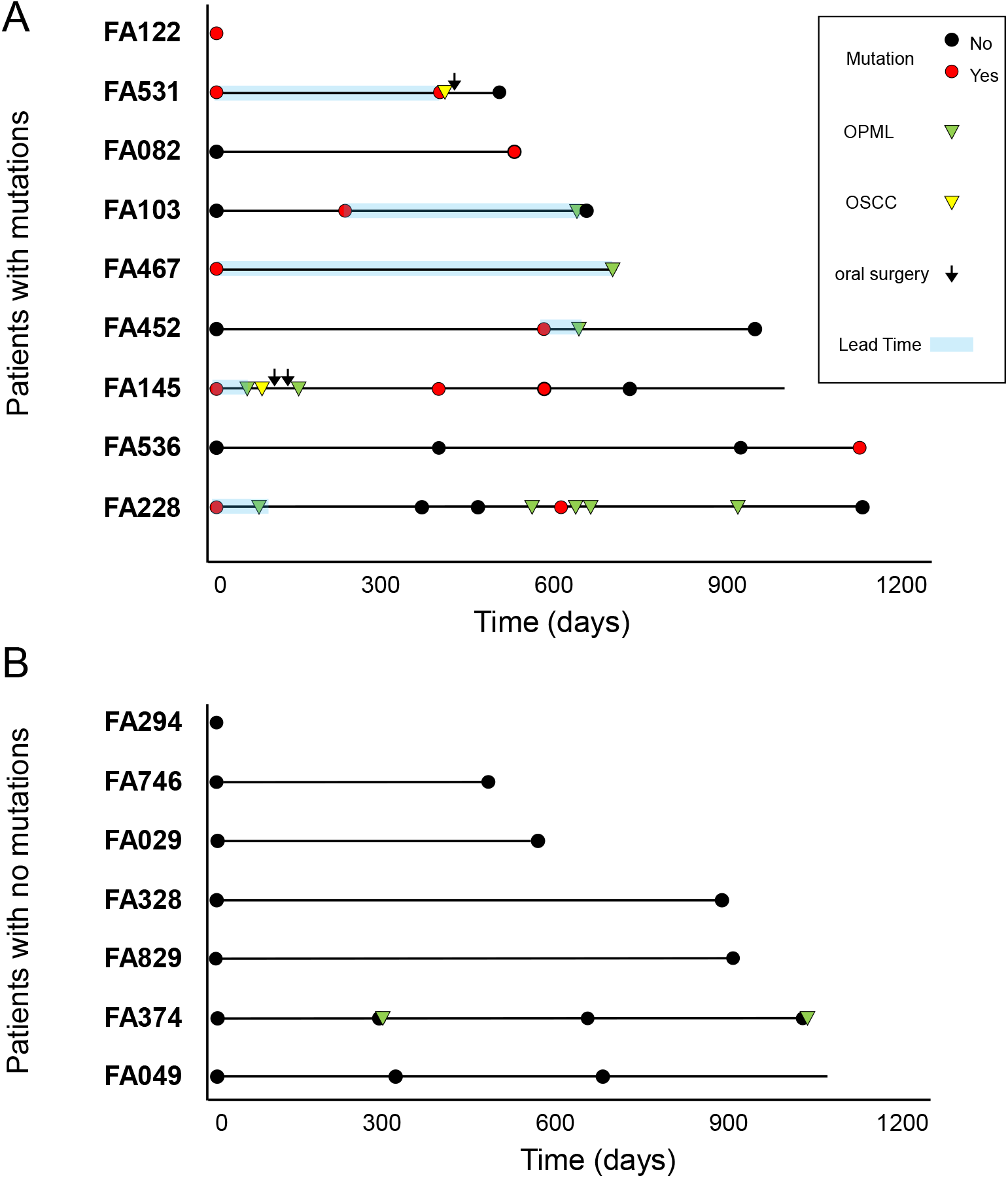
Fanconi anemia patients with mutations develop OPML and/or OSCC. Schematic representation of the molecular and clinical events during patients follow-up. **A)** 9/16 patients had mutations in at least one liquid biopsy sample. 6/9 patients had clinical follow-up data after mutations were found, and all of them displayed OPML or OSCC afterwards. Time from mutation detection to tumor detection (lead time) is shown (blue boxes). Solid horizontal lines represent clinically informed follow-up times from first LB analyzed in each patient. **B)** 7/16 patients displayed no mutations, 6 of whom were free of OPML or OSCC.

We found that 6/7 patients with lesions or tumors had at least one LB sample with a mutation in a cancer gene (Fig. 4A). Importantly, we detected such mutations before clinical diagnosis, with lead times ranging between 23 and 630 days (median: 205 days). On the other hand, we could not find mutations in any of the LB samples from 5 patients free of lesions/tumors (Fig. 4B). Patient FA374 had two leukoplakias during the study, but we could not detect any mutation. No biopsies were taken from these lesions, and therefore no histopathology assessment was conducted (Supplementary Table 5). A Kaplan-Meier curve showed a significant difference in oral lesion/tumor-free survival between FA patients with mutation versus patients with no mutations (Fig. 5A). The accuracy of the diagnostic test to distinguish between patients that will develop oral lesions or carcinomas from those that will not is 92%, with 86% sensitivity and 100% specificity (Fig. 5B). In conclusion, these results showed that the presence of a mutation in saliva or plasma in FA patients is predictive of the appearance of lesions/tumors in the oral cavity region, and can distinguish between high and low risk groups.

**Figure 5.**
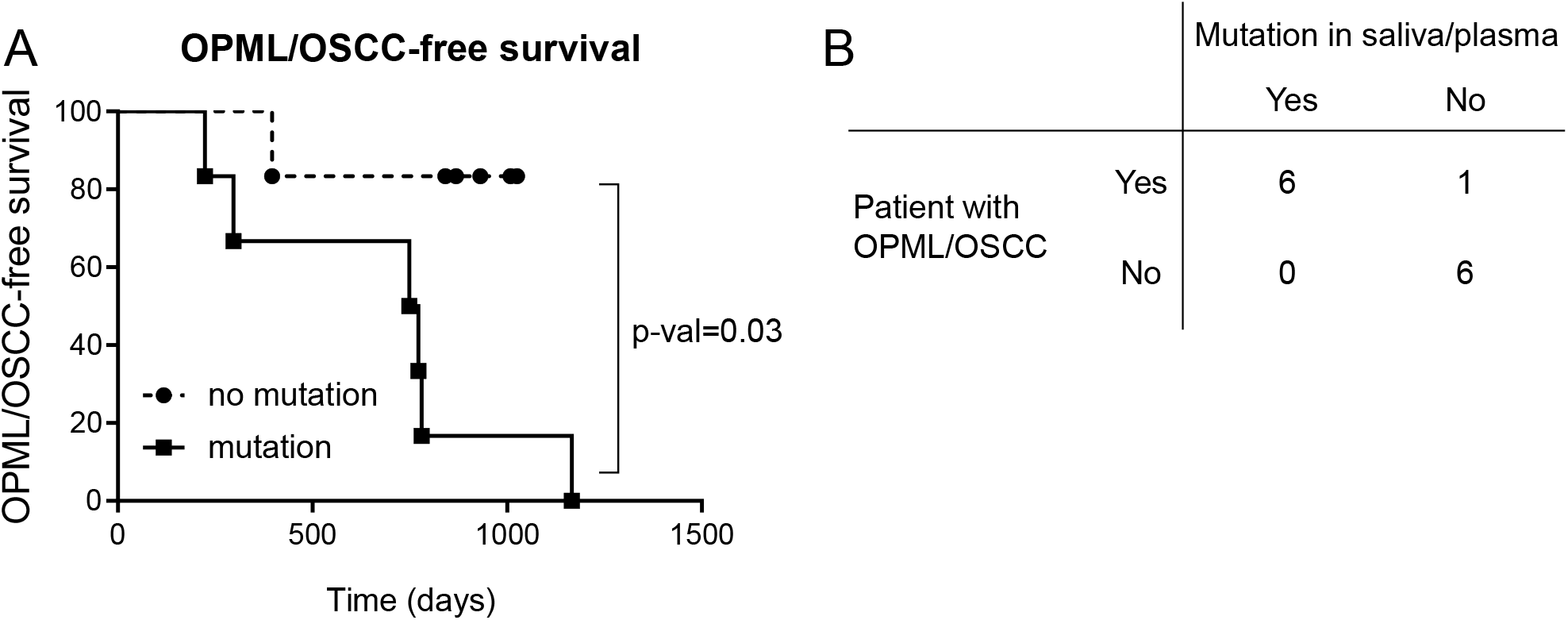
**A)** Kaplan-Meier curves of FA patients with or without mutations. FA patients with mutations display significantly higher risk of developing OPML or OSCC than patients without mutations. Statistical significance differences between curves were determined using log rank test. **B)** Accuracy of the detection of mutations in saliva/plasma as diagnostic for FA patients that develop OPML or OSCC. Global accuracy: 92%; sensitivity: 86%; specificity: 100%.

## DISCUSSION

Mutation analysis of saliva and plasma enabled stratification of Fanconi anemia patients into groups with significantly different risks of developing lesions or squamous carcinomas in the oral cavity. Our proof-of-principle approach involves noninvasive technologies that can be used repeatedly during surveillance of patients to help detect premalignant or malignant cells before clinical symptoms. In addition, other cohorts with high risk of OPMLs and OSCC might benefit from this technology, such as smokeless tobacco users, long-time smokers and heavy alcohol drinkers.

More than 60 years ago, Szilard proposed that accumulation of mutations in somatic cells could accelerate aging in living organisms [24]. Indeed, a number of important reports using sensitive NGS technologies have confirmed the correlation between somatic mutation index and aging in different animal species and organs, such as skin [25] or esophagus [26]. Thus, mutagens such as UV light or tobacco increase mutation rates in cells (in vivo and in vitro) and accelerate senescence, apoptosis, and emergence of cancerous cells. Importantly, most genetic syndromes that impair normal DNA repair mechanisms (such as Fanconi anemia) are associated with premature aging and predisposition to cancer [27]. Importantly, some of these syndromes display high mutational burden in tissues where cancer risk is also higher [28]. Therefore, it is tempting to speculate that tissues from FA patients display exacerbated accumulation of genomic aberrations, which eventually might speed the generation of premalignant and malignant cells. Our results showing that patients with mutations in liquid biopsies are at higher risk of developing oral lesions or squamous carcinomas is compatible with the reported link between increased mutational burden and cancer risk. We hypothesize that some FA patients might have a somatic mutation index higher than other FA patients, and that index could be related with precancer or cancer risk. Future experiments with more sensitive methods and more FA patients in the context of a clinical study should help to validate our hypothesis.

*TP53* is the gene most frequently mutated in OSCC, both in FA patients [23, 29] and in the general population [30]. In addition, *TP53* mutations occur early in the oncogenesis of OSCC, as demonstrated in precancer cell model systems obtained from normal appearing mucosa of the surgical margins of patients with primary OSCCs [31]. Importantly, *TP53* mutations are the most frequent in oral leucoplakias, with higher prevalence in dysplastic versus non-dysplastic lesions [32]. Therefore, the high frequency of *TP53* mutations from early disease stages to invasive carcinomas indicates that such mutations might be found in clinically normal patients using sensitive methods if precancerous or cancerous cells are already existing in the mucosa. Our results indicating the presence of *TP53* mutations in saliva and plasma in patients that developed lesions or carcinomas afterwards suggest that our method can detect such cells before they are clinically visible. Similar results have been reported using NGS in brush biopsies from locations of the oral cavity without visible abnormalities in FA patients. In line with our survival results (Fig. 5A), patients with mutations (frequently in the *TP53* gene) had a higher risk of OSCC than patients with no mutations [32]. Unfortunately, we could not study whether the mutations found in LBs were also present in the consecutive lesion/tumor.

Recent next-generation sequencing analyses have demonstrated that, besides *TP53*, other genes such as CDKN2A and NOTCH1 display small variants both in OSCC tumors and in oral brush biopsies from Fanconi anemia patients [23, 32]. Unfortunately, none of these genes is included in the OCA panel that we used, and therefore we cannot discard the existence of variants in these genes in our LBs. Future implementations of our procedure shall include a gene panel more specific for FA.

In conclusion, deep-sequencing of saliva and plasma from FA patients can detect mutations in cancer genes that stratify FA patients into risk groups. This technology is particularly applicable in FA patients with no visible lesions, such that closer surveillance should be recommended on those displaying such mutations. Further similar longitudinal studies increasing the number of patients and the follow-up time will determine whether our approach allows earlier diagnosis of lesions or carcinomas in the oral cavity.

## Data Availability

All data produced in the present study are available upon reasonable request to the authors

## Acknowledgements

The authors thank J. Bueren, P. Rio, M. Garin, and Jesus M. Paramio for helpful discussions and Norman Feltz for copyreading the manuscript. The authors are also indebted to the patients with FA, their families and clinicians from the Fundacion Anemia de Fanconi (Spain).

