## Supplementary Table 2 for "Early diagnosis of oral cancer and lesions in Fanconi anemia patients: a prospective and longitudinal study using saliva and plasma"

**Supplementary Table 2.** Oncomine™ Pan-Cancer Cell-Free Assay

| Hotspot genes | | | Tumor suppressor genes | CNV genes | Gene fusions * |
|---|---|---|---|---|---|
| *AKT1* | *FGFR3* | *MTOR* | *APC* | *CCND1* | *ALK* |
| *ALK* | *FGFR4* | *NRAS* | *FBXW7* | *CCND2* | *BRAF* |
| *AR* | *FLT3* | *NTRK1* | *PTEN* | *CCND3* | *ERG* |
| *ARAF* | *GNA11* | *NTRK3* | *TP53* | *CDK4* | *ETV1* |
| *BRAF* | *GNAQ* | *PDGFRA* | | *CDK6* | *FGFR1* |
| *CHEK2* | *GNAS* | *PIK3CA* | | *EGFR* | *FGFR2* |
| *CTNNB1* | *HRAS* | *RAF1* | | *ERBB2* | *FGFR3* |
| *DDR2* | *IDH1* | *RET* | | *FGFR1* | *MET* |
| *EGFR* | *IDH2* | *ROS1* | | *FGFR2* | *NTRK1* |
| *ERBB2* | *KIT* | *SF3B1* | | *FGFR3* | *NTRK3* |
| *ERBB3* | *KRAS* | *SMAD4* | | *MET* | *RET* |
| *ESR1* | *MAP2K1* | *SMO* | | *MYC* | *ROS1* |
| *FGFR1* | *MAP2K2* | | | | |
| *FGFR2* | *MET* | | | | |
| • 52 genes<br>• Single library from DNA and RNA*<br>• 272 amplicons<br>• >900 hotspots and indels<br>• Extended coverage of *TP53*<br>• 96 fusions<br>• 12 CNVs<br>• *MET* exon 14 skipping | | | | | |

* RNA was not processed and gene fusions were not interrogated
