## Supplementary Table 3 for "Early diagnosis of oral cancer and lesions in Fanconi anemia patients: a prospective and longitudinal study using saliva and plasma"

**Supplementary Table 3**. Sequencing performance per liquid biopsy sample in FA patients.

| patient | collection time (per patient) | sample type | ng DNA in sample | median molecular coverage[1] | LOD (%)[2] | mapped reads[3] | on target reads (%)[3] |
|---|---|---|---|---|---|---|---|
| FA029 | 1 | plasma | 16.0 | 1508 | 0.2835 | 23,252,734 | 94.5 |
| FA029 | 1 | saliva | 16,500 | 11345 | 0.0767 | 24,820,351 | 96.36 |
| FA029 | 3 | plasma | 21.5 | 1827 | 0.2341 | 11,629,004 | 93.31 |
| FA029 | 3 | saliva | 65,250 | 7718 | 0.077 | 17,646,973 | 96.44 |
| FA049 | 1 | plasma | 18.0 | 970 | 0.4404 | 9,691,979 | 93.4 |
| FA049 | 1 | saliva | 71,250 | 6143 | 0.0771 | 16,753,934 | 97.47 |
| FA049 | 2 | plasma | 10.6 | 1274 | 0.3356 | na | na |
| FA049 | 2 | saliva | 6,213 | 6107 | 0.0771 | na | na |
| FA049 | 3 | saliva | 1,658 | 5863 | 0.0771 | 13,204,407 | 92.92 |
| FA082 | 2 | plasma | 17.7 | 1806 | 0.2368 | na | na |
| FA082 | 2 | saliva | 3,025 | 7576 | 0.077 | na | na |
| FA082 | 4 | plasma | 31.8 | 1103 | 0.3876 | 14,061,179 | 91.42 |
| FA082 | 4 | saliva | 26,500 | 7839 | 0.077 | 17,869,458 | 96.15 |
| FA103 | 1 | plasma | 27.8 | 908 | 0.4704 | 10,700,829 | 94.19 |
| FA103 | 1 | saliva | 35,625 | 8525 | 0.077 | 23,583,046 | 97.29 |
| FA103 | 2 | plasma | 29.5 | 3915 | 0.1093 | 16,066,352 | 94.97 |
| FA103 | 2 | saliva | 12,500 | 11106 | 0.0768 | 22,415,353 | 97.12 |
| FA103 | 3 | saliva | 12,200 | 6576 | 0.0771 | 24,088,967 | 96.99 |
| FA122 | 1 | plasma | 37.1 | 3787 | 0.1129 | 10,544,807 | 92.24 |
| FA122 | 1 | saliva | 24,125 | 7298 | 0.077 | 13,981,295 | 95.81 |
| FA145 | 1 | plasma | 26.5 | 3024 | 0.1414 | 10,997,167 | 93.31 |
| FA145 | 1 | saliva | 34,750 | 5365 | 0.0797 | 10,815,243 | 97.22 |
| FA145 | 2 | plasma | 33.6 | 3334 | 0.1283 | na | na |
| FA145 | 2 | saliva | 11,900 | 5804 | 0.0771 | 23,564,390 | 96.84 |
| FA145 | 3 | saliva | 6,975 | 5339 | 0.0801 | 11,088,276 | 94.8 |
| FA145 | 4 | saliva | 23,125 | 4713 | 0.0908 | 10,415,095 | 94.13 |
| FA145 | 4 | leucocytes | 432 | 3207 | 0.1334 | 12,641,714 | 93.77 |
| FA228 | 1 | plasma | 19.6 | 940 | 0.4545 | 13,408,865 | 93.16 |
| FA228 | 1 | saliva | 27,250 | 12623 | 0.0767 | 25,928,587 | 97.41 |
| FA228 | 2 | saliva | 140,000 | 5836 | 0.0771 | na | na |
| FA228 | 2 | plasma | 9.4 | 957 | 0.4466 | na | na |
| FA228 | 3 | leucocytes | 246.6 | 4085 | 0.1047 | 18,923,515 | 95.63 |
| FA228 | 3 | plasma | 10.7 | 1011 | 0.4228 | na | na |
| FA228 | 3 | saliva | 64,370 | 7992 | 0.077 | na | na |
| FA228 | 4 | plasma | 13.9 | 1622 | 0.2635 | na | na |
| FA228 | 5 | saliva | 52,250 | 7830 | 0.077 | 19,684,271 | 96.84 |
| FA294 | 1 | plasma | 10.4 | 633 | 0.6748 | 9,188,463 | 93.78 |
| FA294 | 1 | saliva | 4,500 | 8605 | 0.077 | 26,520,004 | 97.15 |
| FA328 | 1 | saliva | 7,417 | 4290 | 0.0997 | 12,232,402 | 97.48 |
| FA328 | 1 | plasma | 13.5 | 863 | 0.4952 | 16,046,003 | 93.33 |
| FA328 | 3 | saliva | 1,542 | 4647 | 0.092 | 18,446,600 | 96.01 |
| FA374 | 1 | saliva | 20,375 | 6105 | 0.0771 | 26,463,807 | 97.7 |
| FA374 | 2 | saliva | 6,850 | 13376 | 0.0766 | na | na |
| FA374 | 3 | plasma | 13.6 | 650 | 0.6567 | 9,478,134 | 94.7 |
| FA374 | 3 | saliva | 6,875 | 11185 | 0.0768 | 30,112,655 | 97.21 |
| FA374 | 4 | saliva | 34,250 | 7154 | 0.077 | 19,425,740 | 96.77 |
| FA452 | 1 | saliva | 4,917 | 4637 | 0.0922 | 20,406,290 | 97.72 |
| FA452 | 2 | saliva | 12,500 | 3681 | 0.1162 | 17,321,375 | 95.86 |
| FA452 | 2 | plasma | 11.7 | 548 | 0.7785 | 7,081,164 | 93.4 |
| FA452 | 3 | saliva | 13,375 | 6097 | 0.0771 | 12,785,876 | 94 |
| FA452 | 3 | plasma | 43.8 | 4324 | 0.0989 | 17,938,349 | 92.88 |
| FA467 | 1 | leucocytes | 714 | 3729 | 0.1147 | 19,272,522 | 95.61 |
| FA467 | 1 | plasma | 16.4 | 1859 | 0.23 | na | na |
| FA467 | 1 | saliva | 16,375 | 7868 | 0.077 | na | na |
| FA531 | 2 | saliva | 12,225 | 5072 | 0.0843 | 10,889,921 | 92.08 |
| FA531 | 3 | saliva | 11,775 | 3053 | 0.1401 | 13,829,778 | 94.58 |
| FA531 | 3 | plasma | 10.7 | 1311 | 0.3261 | 16,004,566 | 92.79 |
| FA531 | 4 | saliva | 63,500 | 4103 | 0.1042 | 12,653,106 | 95.91 |
| FA531 | 4 | leucocytes | 285 | 4520 | 0.0946 | 15,931,260 | 94.21 |
| FA536 | 1 | saliva | 13,620 | 7096 | 0.0771 | 36,225,694 | 97.47 |
| FA536 | 1 | plasma | 20.0 | 1079 | 0.396 | 12,515,086 | 92.76 |
| FA536 | 2 | plasma | 15.5 | 1784 | 0.2396 | na | na |
| FA536 | 2 | saliva | 4,875 | 10496 | 0.0769 | na | na |
| FA536 | 3 | saliva | 6,325 | 9233 | 0.0769 | 36,319,049 | 96.87 |
| FA536 | 4 | saliva | 7,925 | 14727 | 0.0765 | 31,401,604 | 97.04 |
| FA746 | 1 | saliva | 788 | 2026 | 0.2111 | 13,898,152 | 95.23 |
| FA746 | 2 | saliva | 3,825 | 6096 | 0.0771 | 30,577,592 | 96.99 |
| FA746 | 2 | plasma | 285.0 | 3247 | 0.1317 | 23,542,906 | 95.81 |
| FA829 | 1 | saliva | 55,000 | 10159 | 0.077 | na | na |
| FA829 | 2 | saliva | 91,250 | 9842 | 0.077 | 29,544,169 | 96.8 |

[1] Median molecular coverage reports median number of individual interrogated DNA molecules across targets.

[2] LOD: limit of detection (median value across all targets)

[3] na: values that could not be retrieved
