## Supplementary Table 4 for "Early diagnosis of oral cancer and lesions in Fanconi anemia patients: a prospective and longitudinal study using saliva and plasma"

**Supplementary Table 4**. Non-synonymous small variants or mutations (SNVs, MNVs and INDELs) found in LBs and leukocytes

| patient | collection time (per patient) | cancer gene | AA change | MAF (%)[1] | saliva | plasma | leukocytes[2] |
|---|---|---|---|---|---|---|---|
| FA082 | 4 | TP53 | p.E180GfsTer67 | 0.1907 | | yes | |
| FA103 | 2 | PDGFRA | p.A466VfsTer39 | 0.0955 | | yes | |
| FA122 | 1 | TP53 | p.R175H | 7.2159 | yes | | |
| FA122 | 1 | TP53 | p.R175H | 33.7177 | | yes | |
| FA145 | 1 | TP53 | p.V157I | 0.0768 | yes | | |
| FA145 | 1 | TP53 | p.W91L | 0.0777 | yes | | |
| FA145 | 1 | TP53 | p.P92A | 0.0932 | yes | | |
| FA145 | 3 | TP53 | p.V157I | 0.0998 | yes | | |
| FA145 | 2 | TP53 | p.V157I | 0.1148 | | yes | |
| FA145 | 4 | PDGFRA | p.A466VfsTer39 | 0.2536 | | | yes |
| FA228 | 1 | TP53 | p.P92A | 0.0837 | yes | | |
| FA228 | 4 | TP53 | p.T102P | 0.2742 | | yes | |
| FA452 | 2 | TP53 | p.W146SfsTer3 | 0.3911 | yes | | |
| FA452 | 2 | TP53 | p.W146SfsTer3 | 0.8229 | | yes | |
| FA467 | 1 | TP53 | p.Q38R | 0.2323 | | yes | |
| FA531 | 2 | TP53 | p.G245S | 0.0726 | yes | | |
| FA531 | 2 | IDH2 | p.R140Q | 0.0758 | yes | | |
| FA531 | 3 | TP53 | p.G245S | 0.2911 | yes | | |
| FA531 | 4 | IDH2 | p.R140Q | 0.1121 | | | yes |
| FA536 | 4 | TP53 | p.P92A | 0.0928 | yes | | |
| FA536 | 4 | TP53 | p.W91L | 0.0986 | yes | | |

[1] MAF: molecular allele frequency

[2] Used to discard possible mutations in saliva/plasma as origitated in white blood cells upon clonal hematopoiesis
