## Supplementary Table 5 for "Early diagnosis of oral cancer and lesions in Fanconi anemia patients: a prospective and longitudinal study using saliva and plasma"

**Supplementary Table 5**. Lesions and SCC during follow-up of FA patients

| Patient | Time order | Location | Oral Lesion | Lesion grade |
|---|---|---|---|---|
| FA145 | 1 | retromolar trigone | leuko/erytroplakia | high grade displasia |
| | 3 | retromolar trigone | margin expansion from SCC | high grade displasia |
| FA452 | 1 | retromolar trigone | leukoplakia | no biopsy |
| FA228 | 1 | palatal mucosa | leukoplakia | low grade displasia |
| | 1 | buccal mucosa | leukoplakia | low grade displasia |
| | 2 | hard palate | leukoplakia | hyperplasia |
| | 2 | buccal mucosa | leuko/erytroplakia | non dysplasia |
| | 3 | right lip | ulcer | hyperplasia/dysplasia |
| | 4 | lip/oral mucosa | leukoplakia | low grade displasia |
| | 5 | vestibular fundus & tongue margin | leukoplakia | hyperplasia |
| FA103 | 1 | tongue | leukoplakia | no biopsy |
| FA467 | 1 | buccal mucosa | leukoplakia | no biopsy |
| FA374 | 1 | alveolar ridge | leukoplakia | no biopsy |
| FA374 | 2 | alveolar ridge & buccal mucosa | leukoplakia | no biopsy |

| Patient | Time order | Location | Oral Cancer | Tumor stage |
|---|---|---|---|---|
| FA531 | 1 | base of tongue | SCC | pT2N0Mx p16- |
| FA145 | 2 | retromolar trigone | SCC | pT1NxMx p16- |
