## Supplementary Table 1 for "Early diagnosis of oral cancer and lesions in Fanconi anemia patients: a prospective and longitudinal study using saliva and plasma"

**Supplementary Table 1.** Fanconi anemia patients' specific characteristics

| Patient | Age[1] | HSCT[2] |
|---|---|---|
| FA122 | 26-30 | no |
| FA531 | 26-30 | no |
| FA82 | 46-50 | no |
| FA103 | 26-30 | yes |
| FA467 | 11-15 | yes |
| FA452 | 16-20 | yes |
| FA145 | 21-25 | yes |
| FA536 | 16-20 | yes |
| FA228 | 31-35 | yes |
| FA294 | 36-40 | yes |
| FA746 | 16-20 | yes |
| FA29 | 26-30 | no |
| FA328 | 16-20 | yes |
| FS829 | 36-40 | no |
| FA374 | 16-20 | yes |
| FA49 | 26-30 | yes |

[1] Age at patient recruitment
[2] HSCT: hematopoietic stem cell transplantation
